# SARS-CoV-2 receptor ACE2 is expressed in human conjunctival tissue, especially in diseased conjunctival tissue

**DOI:** 10.1101/2020.05.21.20109652

**Authors:** Shengjie Li, Danhui Li, Jianchen Fang, Qiang Liu, Wenjun Cao, Xinghuai Sun, Gezhi Xu

## Abstract

COVID-19 virus has currently caused major outbreaks worldwide. ACE2 is a major cellular-entry receptor for the COVID-19 virus. Although ACE2 is known to be expressed in many organs, whether it is expressed by the conjunctival tissue is largely unknown. Human conjunctival tissues from 68 subjects were obtained, which included 10 subjects with conjunctival nevi, 20 subjects with conjunctivitis, 9 subjects with conjunctival papilloma, 16 subjects with conjunctival cyst, 7 subjects with conjunctival polyps, and 6 ocular traumas as normal subjects. Expression of ACE2 was evaluated by immunohistochemistry, immunofluorescence, reverse transcriptase-quantitative polymerase chain reaction, and western blot assay. We observed the expression of ACE2 by conjunctival tissues, expecially in conjunctival epithelial cells. ACE2 was significantly (p<0.001) overexpressed in conjunctival cells obtained from subjects with conjunctivitis, conjunctival nevi, conjunctival papilloma, conjunctival cyst, and conjunctival polyps epithelial cells when compared to that in conjunctival epithelial cells obtained from control subjects. Collectively, clinical features of reported COVID-19 patients combined with our results indicate that COVID-19 is likely to be transmitted through the conjunctiva.

## To the Editor

In December 2019, coronavirus disease 2019 (COVID-19) was first reported in people in Wuhan, China, and has rapidly spread throughout the world [1]. As of August 11, 2020, 19,936,210 laboratory-confirmed cases were reported. Furthermore, the World Health Organization reported 732,499 fatalities [2]. The typical signs and symptoms of COVID-19 are fever, dry cough, and fatigue. The virus is mainly transmitted through respiratory droplets and by close contact with infected individuals. Furthermore, about 2.78% of COVID-19 patients showed symptoms of conjunctival congestion [3]. Thus, because the ocular surface is always exposed to the environment, it may serve as a possible site of virus entry and also as a source of contagious infection.

Studies have provided powerful evidence that angiotensin-converting enzyme 2 (ACE2) is the major cellular-entry receptor for the SARS-CoV-2 virus, and higher ACE2 expression increases susceptibility to SARS-CoV-2 [4]. Numerous studies, along with the typical symptoms exhibited by COVID-19 patients [3,5], suggest that the expression of the *ACE2* gene may be a factor involved in the mode of transmission of the virus. Recently, Collin et al. [6] and Zhou et al. [7] reported that ACE2 was expressed in normal human adult conjunctival, limbal and corneal epithelium, suggesting that ocular surface cells could be susceptible to infection by SARS-CoV-2. These initial reports led us to question whether and how the *ACE2* gene is expressed in diseased conjunctival tissues.

In the current study, we analysed both normal and diseased human conjunctival tissue using RT-qPCR, Western blot, immunohistochemistry, and immunofluorescence in order to determine both expression and localization of this key viral susceptibility factor in the human conjunctival tissue. Sixty-eight subjects with a mean age of 47.42 ± 16.19 years [(32 males (47.06%); 36 females (52.94%)] were recruited for the study, which included 10 subjects with conjunctival nevi, 20 subjects with conjunctivitis, 9 subjects with conjunctival papilloma, 16 subjects with conjunctival cyst, and 7 subjects with conjunctival polyps; 6 ocular traumas were used as normal control subjects. More information related to materials and methods can be found in online supplementary materials. All analyses were performed using the Statistical Package for the Social Sciences software, version 13.0 (SPSS Inc., Chicago, IL, USA).

As presented in Fig. 1A and B, ACE2 mRNA and protein levels were significantly overexpressed in conjunctival tissues of inflamed conjunctiva, conjunctival nevi, conjunctival cyst, conjunctival papilloma, and conjunctival polyps, as compared to normal conjunctival tissues (p < 0.001). Furthermore, a significant difference in ACE2 mRNA and protein levels between normal tissue and diseased conjunctival tissues was observed (p < 0.001; Fig. 1A and B).

**Fig. 1.**
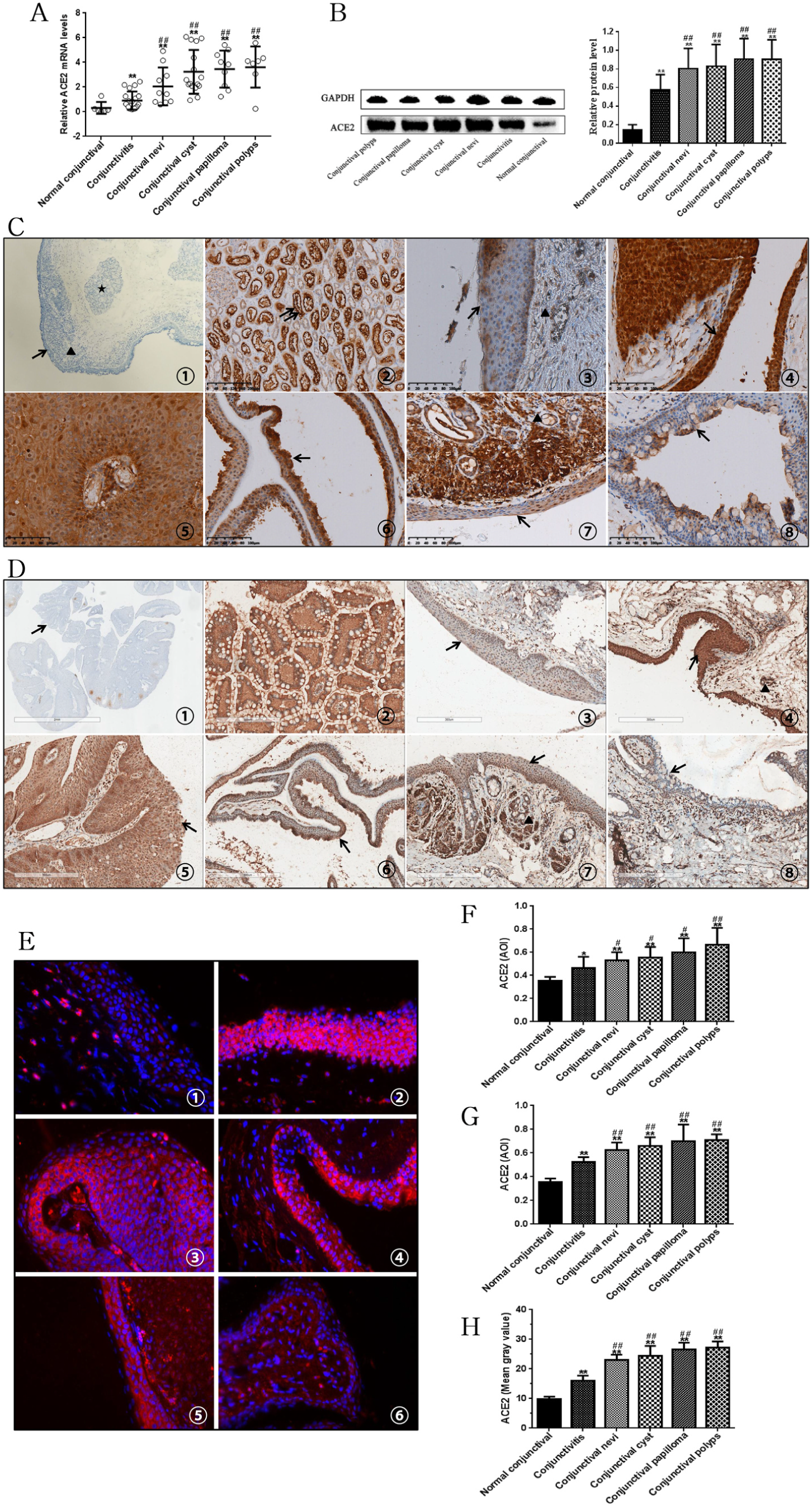
ACE2 mRNA levels (A) and protein levels (B) in different type conjunctival tissues were examined by RT-qPCR and Western blot assay. Polyclonal rabbit anti-ACE2 antibody (C) and monoclonal mouse anti-ACE2 antibody (D) were used in immunohistochemistry. Negative controls used PBS instead of primary antibodies (C, D). The kidney was used as positive control (C②, D②) and kidney tubules were marked with double arrows. Normal conjunctival tissue (C③, D③). Strong staining is present in conjunctival polypus (C④, D④), conjunctival papilloma (C⑤, D⑤), conjunctival cyst (C⑥, D⑥), and conjunctival nevi (C⑦, D⑦). Expression of ACE2 was weakly positive in conjunctivitis (C⑧, D⑧). The normal conjunctiva consists of the epithelium (arrow) and stroma (triangle). The stroma is located under the epithelium and is a loose fibrous vascular tissue containing accessory larcimal glands (star), lymphocytes and nerves. Immunoreactivity is denoted by red staining (E, DAPI: blue). Normal conjunctival tissue: E. Granular ACE2 staining is present in the epithelium. Strong staining is present in conjunctival polypus (E②), conjunctival papilloma (E③), conjunctival cyst (E④), and conjunctival nevi (E⑤). Result expression of ACE2 was weakly positive in conjunctivitis (E⑥). Magnification 40x. Summary of the quantitative immunohistochemical (F, polyclonal rabbit anti-ACE2 antibody; G, monoclonal mouse anti-ACE2 antibody) and immunofluorescence (H) stainingTop of the box plot represents the mean; the bar of each box represents the standard deviation. Mann–Whitney *U* test was used. *P < 0.05, **P < 0.001 vs. the normal conjunctival tissue.^#^P < 0.05, ^##^P < 0.001 vs. conjunctival tissues of inflamed conjunctiva.. (For interpretation of the references to colour in this figure legend, the reader is referred to the Web version of this article.)

Two different antibodies, polyclonal rabbit anti-ACE2 antibody (Fig. 1C) and monoclonal mouse anti-ACE2 antibody (Fig. 1D), were used for immunohistochemistry. Notably, we observed the presence of ACE2 in normal conjunctival epithelial cells (Fig. 1C③, Fig. 1D③). Marked ACE2 immunostaining was also found in epithelial cells in tissue samples from conjunctival polyps (Fig. 1C④, Fig. 1D④), conjunctival papilloma (Fig. 1C⑤, Fig. 1D⑤), conjunctival cyst (Fig. 1C⑥, Fig. 1D⑥), conjunctival nevi (Fig. 1C⑦, Fig. 1D⑦), and conjunctivitis (Fig. 1C⑧, Fig. 1D⑧). Marked ACE2 immunofluorescence staining was also found in samples from normal conjunctival tissue (Fig. 1E), conjunctival polyps (Fig. 1E②), conjunctival papilloma (Fig. 1E③), conjunctival cyst (Fig. 1E④), conjunctival nevi (Fig. 1E⑤), and conjunctivitis (Fig. 1E⑥). The levels of ACE2 staining were significantly different between diseased and normal conjunctival epithelial cells.

A summary of the quantitative data is presented in Fig. 1F–H. Significant differences in ACE2 expression between normal and diseased conjunctival epithelial cells were observed (p < 0.05). Among diseased conjunctival tissues, the level of ACE2 was significantly (p < 0.001) lower in inflamed conjunctiva as compared to other diseased tissue. The most significant finding was the surface expression of ACE2 in conjunctival epithelial cells. Recently, Collin et al. [6] reported that ACE2 was expressed in the human adult conjunctival, limbal and corneal epithelium. Meanwhile, Zhou et al. [7] also found ocular surface expression of ACE2 proteins across all human ocular specimens. Our results confirm these reports, but go further in showing that ACE2 was overexpressed in diseased tissue. Several respiratory viruses have been shown capable of using the eye as both a site for replication and a port of entry, which could result in a productive respiratory infection [8]. Peiris JS et al. [9] reported that SARS-CoV can be transmitted either through Please cite this article as: Shengjie Li, *The Ocular Surface*, https://doi.org/10.1016/j.jtos.2020.09.010 direct or indirect contact with ocular mucous membranes. Collectively, it is evident that the virus, via droplets, might be transmitted through the human conjunctival epithelium.

Our study shows that ACE2 was significantly overexpressed in diseased epithelial cells when compared to normal conjunctival epithelial cells. Collin et al. [6] suggested that local inflammation can enhance ACE2 expression, which may explain why ACE2 is expressed at higher levels in diseased conjunctiva. Guangfa Wang, a member of the national expert panel on pneumonia, was infected by the COVID-19 virus during an inspection in Wuhan. Wang complained of redness of the eyes, which suggests that he was infected through the ocular surface [10]. Xia J et al. [11] reported that conjunctival swab samples collected from patients with conjunctivitis yielded positive SARS-CoV-2 RT-qPCR results, whereas negative RT-qPCR results were obtained in patients without conjunctivitis. One explanation for why patients with conjunctivitis are more likely to exhibit positive SARS-CoV-2 RNA levels is that conjunctivitis may have resulted from the active presence of SARS-CoV-2. On the other hand, the subjects with conjunctivitis may have had higher levels of ACE2, which may lead to a much stronger susceptibility to SARS-CoV-2 infection.

Our results show that ACE2 is present in human conjunctival tissue, especially in diseased conjunctival tissue. Collectively, the clinical features of reported COVID-19 patients combined with our results indicate that COVID-19 is likely to be transmissible through the conjunctiva. It is easy to forget that conjunctivae are generally exposed to the environment. Thus, increasing the awareness of eye protection during COVID-19 is necessary, especially for subjects with diseased conjunctivae.

## Data Availability

All data has been shown in manuscript.

## Ethical statement

The ethics committee of the Renji Hospital of Shanghai Jiao Tong University approved this study, and it adhered to the principles of the Declaration of Helsinki. Informed consent was obtained from all participating subjects.

## Acknowledgements

This work was supported by Shanghai Sailing Program (18YF1403500), Shanghai Municipal Commission of Health and Family Planning (20174Y0169), Shanghai Municipal Commission of Health and Family Planning (201840050), Shanghai Science Technology Committee Foundation grant (19411964600), the State Key Program of National Natural Science Foundation of China (81430007), the subject of major projects of National Natural Science Foundation of China (81790641), the Shanghai Committee of Science and Technology of China (17410712500), and the top priority of clinical medicine center of Shanghai (2017ZZ01020).

## Appendix A. Supplementary data

Supplementary data to this article can be found online at https://doi.org/10.1016/j.jtos.2020.09.010.

